# SARS-CoV-2 Omicron disease burden in Australia following border reopening: a modelling analysis

**DOI:** 10.1101/2022.03.09.22272170

**Authors:** George J Milne, Julian Carrivick

**Affiliations:** Department of Computer Science and Software Engineering and The Marshall Centre for Infectious Diseases, University of Western Australia, Perth, Australia

## Abstract

**Background:** Countries with high COVID-19 vaccination rates have seen the SARS-CoV-2 Omicron variant result in rapidly increasing case numbers. This study evaluated the impact on the health system which may occur following introduction of the Omicron variant into Western Australia following state border reopening. We aimed to understand the effect of high vaccine coverage levels on the population health burden in the context of lower vaccine effectiveness against the Omicron variant, the impact of a third dose booster regime, and ongoing waning of vaccine-induced immunity. Originally scheduled for 5^th^ February 2022, the Western Australian border was opened on 3^rd^ March 2022, we also aimed to determine the impact of delaying border reopening on the COVID-19 health burden and whether the West Australian health system would be able to manage the resulting peak demand.

**Methods:** An agent-based model was used to evaluate changes in the COVID-19 health burden resulting from different border openings, at monthly intervals. We assumed immunity was derived from vaccination with the BNT162b2 Pfizer BioNTech vaccine and waned at observed rates from the UK. The model was calibrated against outbreaks in two other Australian states, Queensland and South Australia, both of which were in a similar situation to Western Australia with negligible COVID-19 transmission prior to Omicron’s introduction. Age-specific infections generated by the model, together with recent UK data, permitted resulting outbreak health burden to be quantified, in particular peak ICU demand.

**Results:** Overall population immunity in Western Australia is shown to peak and then plateau for a period of 5 months, between February and June 2022, resulting in a similar health burden if the border is reopened prior to June 2022. For an opening date of 5^th^ March 2022, hospitalisations are predicated to peak at 510 beds, 51 of which will be in ICU, with a total of 383 deaths. If the border reopened on 5^th^ June 2022, hospitalisations are expected to peak with 750 beds required, 75 of which would be in ICU, and a total of 478 deaths. With a total surge capacity of 52 fully staffed ICU beds, West Australian hospitals are predicted to have adequate ICU capacity for future COVID-19 demands if border reopening occurs prior to May 2022.

**Conclusions:** Our results show that with extremely high SARS-CoV-2 vaccination rates in Western Australian, and documented vaccine-induced vaccine waning rates, the overall population immunity in Western Australia will be at its highest in the period of February 2022 to June 2022. Opening the Western Australian border prior to the end this period will result in the lowest health burden in comparison to opening in June 2022 or later. With a border reopening of 3^rd^ March 2022 announced by the Western Australian government, our data for a 5^th^ March 2022 opening date may be used to predict the progression of this resulting outbreak. These data show expected peak demand of 510 hospital beds, 51 of which will be in ICU, with a total of 383 deaths. With a surge capacity of 52 ICU beds, it is expected that the Western Australian hospital system will be able to handle the additional load during the peak of the wave.

## Background

Countries with high COVID-19 vaccination rates have seen the SARS-CoV-2 Omicron variant result in rapidly increasing case numbers. This study evaluated the impact on the health system which may occur following introduction of the Omicron variant into Western Australia following state border reopening. We aimed to understand the potential effectiveness of high vaccine coverage levels in the following context: lower vaccine effectiveness against the Omicron variant; the impact of a third booster vaccination regimen; and ongoing waning of vaccine-induced immunity. Until February 2022 there was limited SARS-CoV-2 transmission in Western Australia due to strict international and inter-state border controls. This resulted in the population having almost no immunity due to prior infection.

The aim of this study was to predict the increased hospital demand resulting from ongoing Omicron transmission, particularly the impact on Intensive Care Units, once the state border reopens. Unlike other Australian states, Western Australia retained semi-closed borders, requiring quarantine measures for those permitted to enter. A 5^th^ February 2022 border opening date for Western Australia was delayed, and the Government of Western Australia subsequently announced that strict border controls for incoming travellers would cease on 3^rd^ March 2022.^1^ A key aim of this study was to determine how delaying border opening, and thus on-going Omicron variant transmission, impacts the resulting Omicron health burden.

From November 2021 onwards the highly transmissible SARS-CoV-2 Omicron variant spread worldwide, including in countries with high vaccination rates.^2,3^ Omicron has been shown to be more transmissible than the Delta variant but less pathogenic, resulting in a rapid growth in case numbers but a lower number of hospitalisations and deaths than seen with Delta.^4^ Significantly, a two-dose vaccination regimen has been shown to be less effective in preventing Omicron infection and severe disease when compared to Delta.^4^ However, booster third doses of mRNA vaccines have been found to be highly effective.^4^

## Methods

### The UWA COVID-19 model

An individual-based model capturing the demographics and movement patterns of individuals within the Australian city of Newcastle (population ∼273,000), together with SARS-CoV-2 virus transmission data from Wuhan, China prior to social distancing activation,^5^ was previously developed (February 2020). That model was used to analyse the effectiveness of non-pharmaceutical social distancing interventions, varying their stringency, timing, and duration.^6,7^ Our model was re-parameterised to reflect the transmission characteristics of the Delta variant following its emergence, and used to analyse a wide range of alternative vaccination strategies (October 2021).^8^ These highlighted the importance of vaccinating children and adolescents, and determined vaccine coverage levels needed to reduce and prevent growth in case numbers, with and without reintroduction of social distancing measures. The rationale for using a high-resolution agent/individual type model is its ability to determine the ages of those infected under alternative vaccination strategies. This permits estimation of (inherently) age-specific health burden outcomes occurring during future COVID-19 epidemics, such as that resulting from introduction of the Omicron variant into a population. The potential of alternative response measures to mitigate the resulting health burden may thus be quantified as reductions in cases, hospitalisations, ICU demand and deaths.

### Simulation algorithm

As described in prior studies,^6-9^ our model represents every individual in the modelled community, together with their age. Each individual has an associated data structure to track their infection state (*i.e*. SEIR) which is updated in response to simulation events, such as virus transmission and waning immunity. Individuals are explicitly moved between households, schools and workplaces in each 12-hour cycle, in which interaction may occur with other individuals. These interactions are pairwise, where the *probability of transmission* between an infected and susceptible individual is governed by a transmission parameter, called *beta*. This parameter is specific to the SARS-CoV-19 variant being modelled, and relates to its basic reproduction number.^10^ A given *beta* is obtained by infecting a single random individual in the model, with all other individuals in the susceptible state. The model software is then run and the number of secondary infections which occur are logged, with the process repeated multiple times to overcome stochastic uncertainty. The resulting secondary infection distribution provides the mean population-wide basic reproduction number R_0_ resulting from a given *beta* setting. This process was conducted repeatedly, adjusting the *beta* up or down until a target reproduction number was obtained.

The model captures pathogen transmission and thus generates day-to-day infection data. This data includes individuals who may never get detected as a diagnosed case due to asymptomatic infections, or imperfect testing compliance. Infection data is therefore converted into diagnosed cases using a 2:1 ratio, as used in a prior study and which reflects estimated COVID-19 infection/case ratios in Australia.^8,11^

### Omicron variant

We estimated an effective reproduction number R_eff_ excluding any immunity for Omicron of 3.9 using daily hospitalisation data from two Australian states: Queensland and South Australia.^19^ COVID-19 hospitalisation data was used as this was determined to better reflect the rate of Omicron transmission within the community compared to reported daily case data. Once borders reopened in these states and Omicron transmission increased rapidly from December 2021 onwards, PCR testing facilities became overwhelmed, and testing compliance declined.^20^ This feature is highlighted in Figure 2, where observed and predicted cases rapidly decouple after approximately 3,000 daily cases are reached. However, fitted and observed daily hospitalisations are seen to be well matched.

**Figure 1.**
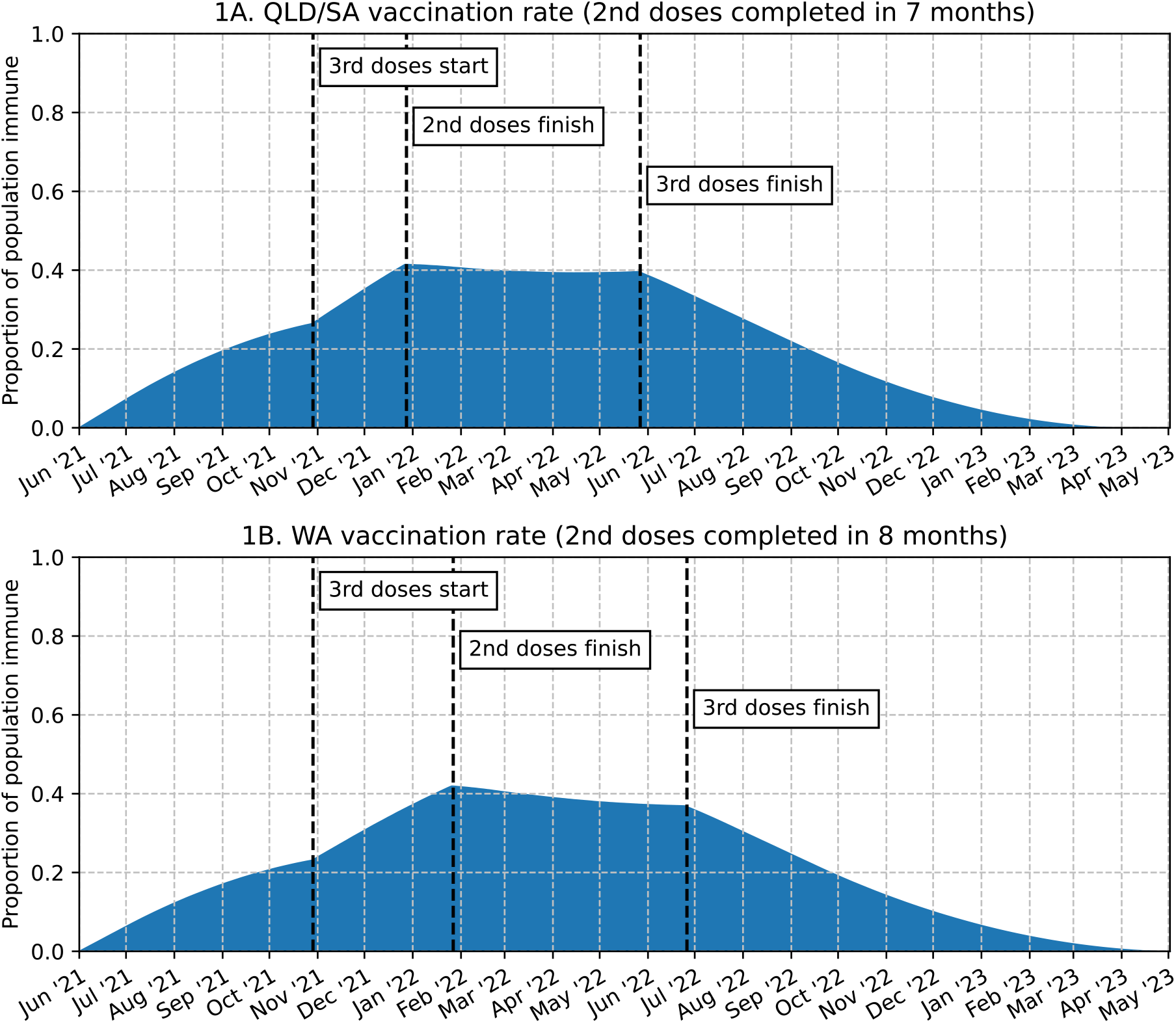
Population immunity dynamics due to vaccination. Second doses administered at rate to reach 90% coverage of ages 12+ over 7 and 8 months. Third doses administered 5 months after second dose. Third dose vaccinations completed after ∼12 months.

**Figure 2.**
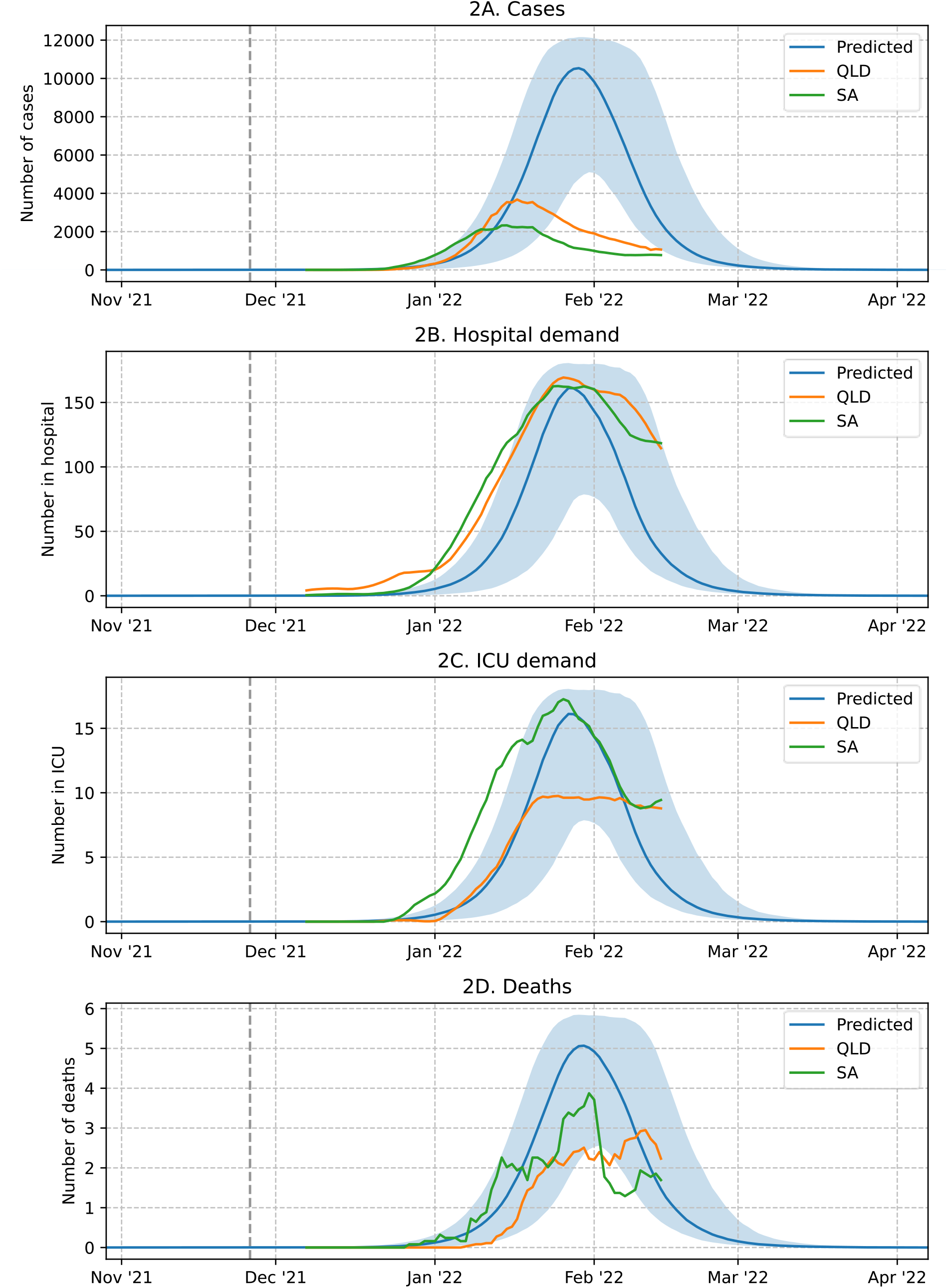
7 day moving average of actual and predicated daily cases, hospitalisations, ICU demand and fatalities per million after fitting to Queensland and South Australia hospitalisation data.

Published UK data on vaccine effectiveness against Omicron variant infection and severe illness has been used in our Omicron model.^4,18^ This data provided estimates of the effectiveness of second and third booster doses of the BNT162b2 Pfizer vaccine, and the rate of waning vaccine effectiveness to both of these doses. With an approximate delay of around 5 months between second and third doses,^15^ this permitted the time-changing *immunity profile* in the Queensland, South Australian and Western Australian populations to be determined in the absence of Omicron transmission, for the state-specific second dose vaccination rates illustrated in Figure 1. Different second dose (and third dose) vaccination rates can be seen to affect the time changing immunity profile of the overall population. In Queensland and South Australia, whose Omicron hospitalisation data was used in the model calibration process, second doses were administered over a 7 month period.^15^ In Western Australia, second doses were administered at a slower rate, over an 8 month period.^15^ A comparison of the population immunity derived from each vaccination scenario appears in Figure 1.

Importantly, the Omicron immunity profile for Western Australia captures three concurrent dynamical processes, *viz. i)* second dose vaccinations; *ii)* third dose vaccinations; *iii)* and the waning of vaccine-induced immunity. These dynamics are key to modelling the health burden resulting from alternative Western Australia border opening dates, with data presented in Figure 3 and associated tables.

**Figure 3.**
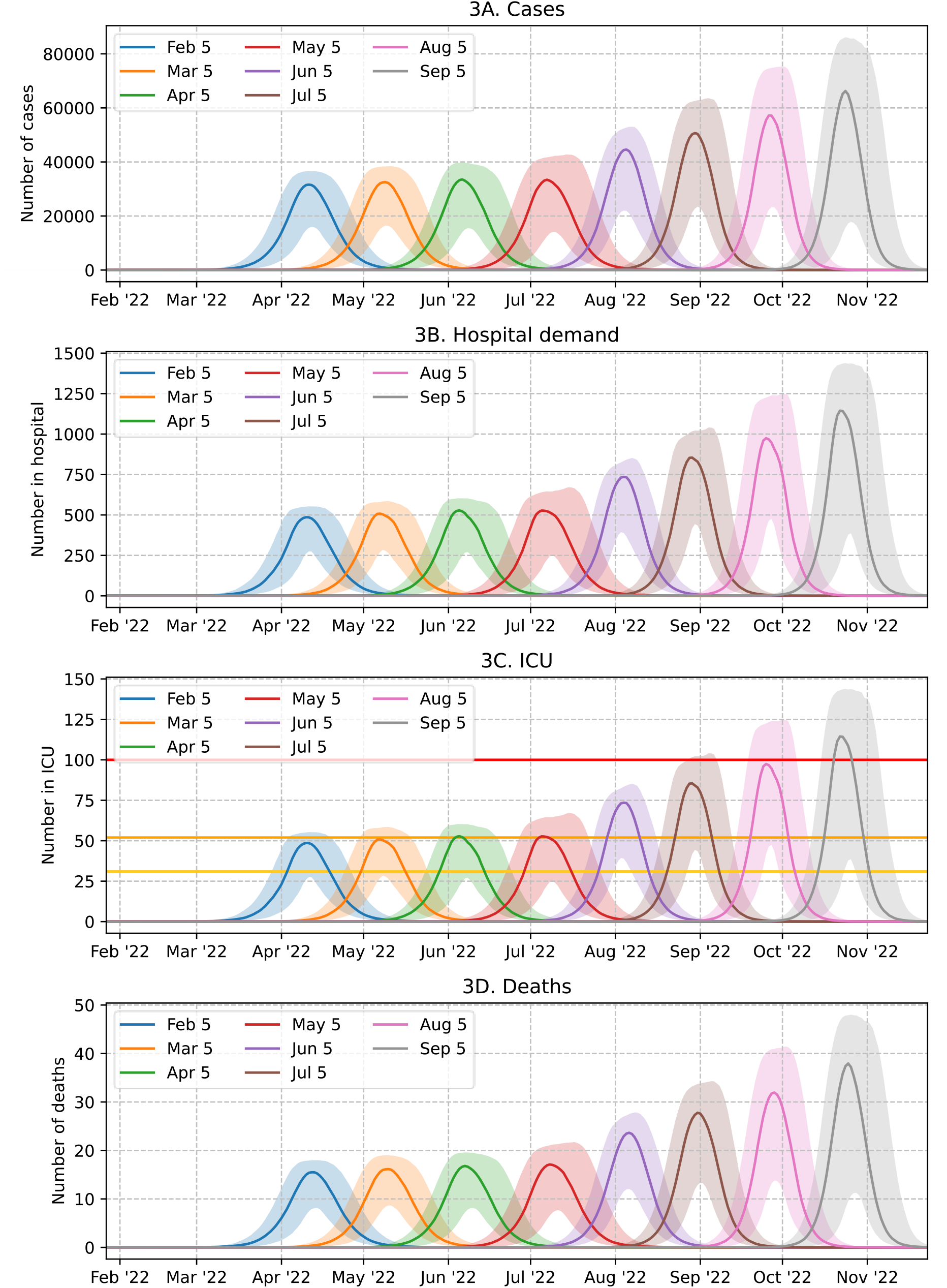
7 day moving average of predicted daily cases, hospitalisations, ICU demand and fatalities for Omicron epidemic in Western Australia, for alternative border opening dates, for population of 2.7 million persons. In Figure 3C, horizontal lines represent ICU bed thresholds: 31 currently available fully-staffed excess bed capacity (yellow), 52 surge (orange) and 100 (red) beds, as described by Litton, Huckson ^30^.

### Vaccination

We modelled the BNT162b2 Pfizer vaccine with assumed protection against Omicron consistent with UK observations^4^ and of second and third doses administered 5 months apart.^15^ In the absence of further evidence we assumed protection against infection to be the same as protection against disease. In the model the effect of vaccination is captured at the individual level, where a certain percentage of fully vaccinated individuals “move” from a susceptible state to an immune state, as in a classical SEIR approach. The percentage of individuals moving to an immune state thus reflects vaccine efficacy. Using this, the total population immunity from vaccination can be represented, and thus the time-changing immunity profile of the overall modelled population due to vaccination may be represented over time. This immunity dynamics is implemented in the simulation software, together with daily pairwise SARS-CoV-19 virus transmission, and the effect of waning vaccine-induced immunity.

The study assumed a setting applying to certain Australian states prior to introduction of the Omicron variant, with effectively no SARS-CoV-2 transmission and thus no immunity due to prior infection. The states of South Australia, Queensland, Tasmania, and the Northern Territory, had minimal SARS-CoV-2 transmission due to early international and inter-state border closures and adoption of a SARS-CoV-2 elimination policy.^21^ We thus assume all immunity effects are attained as the result of vaccination, rather than immunity derived from infection.

Australia adopted an age-specific vaccination strategy in March 2021 due to limited supplies of mRNA vaccines, with those aged 60 year and over vaccinated with the AstraZeneca vaccine, manufactured locally, with all younger age groups vaccinated with the Pfizer vaccine. Our study assumed a modified vaccination strategy by using the Pfizer mRNA vaccine to boost the immune response of those previously vaccinated with the AstraZeneca vaccine, found necessary due to its lower efficacy against the Omicron variant.^4^ This simplified the modelled scenarios by assuming all vaccinated individuals had either a second or third dose of the Pfizer vaccine, and thus the same protective effect following their most recent vaccination. From October 2021 Australia introduced the Pfizer vaccine to boost immunity levels for those vaccinated 6 months previously with either the Astra Zeneca or Pfizer vaccines.^22^

Protection against poor health outcomes in those vaccinated and having breakthrough infections was implicitly included by using recent UK Omicron variant data to give age-specific case/hospitalisation and case/mortality ratios, as detailed in Table S1.^19^ Further model parameters are presented in Table 1.

**Table 1:** Omicron variant parameter settings

|  |  |
| --- | --- |
| **Model population** | 273,407. 2011 census data for Newcastle and Lake Macquarie East, NSW. |
| **R<sub>eff</sub> excluding immunity** | 3.9 ([3.8,4.0] 95% CI) |
| **Time from infection to symptoms** | 3 days<sup>12</sup> |
| **Time from infection to infectious** | 2 days |
| **Time from infection to recovery** | 8 days (5 days of symptoms, 6 days being infectious) |
| **Child (<12 years) susceptibility** | Same as adults & adolescents |
| **Child (<12 years) transmissibility** | 50% of adults |
| **Adolescent (12-17 years) transmissibility** | Same as adults |
| **Probability of asymptomatic infections** | 35%<sup>13</sup> |
| **Probability of asymptomatic transmission** | 55% of symptomatic transmission<sup>14</sup> |
| **Probability of symptomatic infection isolation** | 33% |
| **Home isolation of diagnosed cases** | 7 days |
| **Seeded infections** | On average 1 infection every 4 days for 30 days |
| **BNT162b2 Pfizer vaccine** | Efficacy: Matching UK data<sup>4</sup><br>second doses administered at the required rate to reach 90% of ages 12+ in:<br>• Fitting: 7 months (QLD/SA)<sup>15</sup><br>• Projecting: 8 months (WA)<sup>15</sup><br>Third doses administered 5 months after second dose<sup>15</sup> |
| **Length of Hospital Stay** | 4 days<sup>16,17</sup> |
| **Infection/Case Ratio** | 2:1<sup>11</sup> |
| **Case/Hospitalisation Ratio**<br>**Case/Fatality Ratio** | Matching age specific UK data<sup>18</sup> |
| **Hospitalisations/ICU Ratio** | 10:1<sup>19</sup> |

### Waning immunity

Waning immunity was captured by altering the immune state of individuals whose immunity resulted from vaccination, returning their immune state back to a susceptible state, at a waning rate taken from UK vaccine effectiveness data.^4^ This data indicates that a BNT162b2 Pfizer second dose provides 65% of individuals with immunity for 30 days, before waning to 15% over a further 140 days. Prior to completion of this waning period (after 5 months) a third dose is assumed to be administered, returning a certain percentage of those who received the boosting dose to an immune state, so boosting the overall population immunity profile, as seen in Figure 1. This results in 70% of vaccinated individuals regaining immunity, the effect of the third boosting dose is then assumed to wane after 28 days, and the waning cycle repeats itself, with all individuals losing their protection after a further 294 days, a period extrapolated from the limited post third dose data.^4^

We assume that individuals who recover from an Omicron infection have immunity from reinfection for the duration of the simulation period. While data is still emerging regarding the protection gained from Omicron infection against future Omicron reinfection, prior studies comparing Alpha, Beta and Delta reinfection suggest that protection level resulting from infection to be greater than 90% in each of these variants.^23^ In the absence of further evidence, we assume this long-term immunity will also hold as a consequence of Omicron infection. As reinfections are defined as two positive PCR tests more than 90 days apart,^23^ and this time period is longer than the generated epidemic curves, any effects of reinfection risk are unlikely to affect our results.

### Model calibration

Transmission parameters in the model were calibrated using specific Omicron variant outbreak data from two Australian states, outbreaks which occurred following border reopening in November 2021.^24,25^ Data from the states of Queensland and South Australia were used, given *1)* similar vaccination coverage rates to Western Australia, and *2)* limited SARS-CoV-2 transmission prior to the arrival of Omicron, a similar situation to Western Australia on border reopening. Hospitalisation data were used to fit model parameters to actual Omicron outbreak data, rather than case number data. This was due to the breakdown of testing, and thus the accuracy of case numbers, once Omicron outbreaks gained significant momentum, when case numbers began to exceed testing capacity.^20^

As explained previously, the *beta* transmission coefficient, which models the inherent transmissibility of the Omicron variant, was determined experimentally, giving an effective reproduction number R_eff_. This reflected the Omicron transmission characteristics in the South Australia and Queensland populations where limited non-pharmaceutical measures (NPI) were in place, such as restricted densities and wearing masks in public indoor spaces, but with no population immunity from vaccination (which was applied during the calibration procedure, and would reduce this value depending on the time-specific population immunity shown in Figure 1). They also included observed self-activated NPIs, such as working from home.^26^ These measures are assumed to occur in Western Australia when Omicron case numbers increase and borders reopen.

The calibration procedure proceeded with an initial *beta* value, and assumed a second dose vaccination rollout period of 7 months, as occurred in Queensland and South Australia (Figure 1A),^15^ starting on the 1^st^ June 2021. Infectious cases are seeded into the model after 180 days (early December) to model the significant Omicron case numbers which began to appear in those states following their borders opening.^19^ The *beta* transmission parameter was adjusted repeatedly in a series of simulation experiments to generate daily Omicron infection data, and UK case/hospitalisation ratios were applied to determine hospitalisation outcomes.^18^ This fitting procedure was repeated, adjusting the *beta* parameter until hospitalisation patterns matched the 7 day moving hospitalisation average reported for Omicron outbreaks in Queensland and South Australia.^19^ This process resulted in an effective reproduction number R_eff_ of 3.9.

### Health burden outcomes

The model captures the age of each modelled individual in one of 10 age bands, allowing age-specific health outcomes to be determined following infection.^9^ Model outputs capture the infection history of all individuals in the community, representing where and when infections occur. The modelling analyses quantified infection and case reductions arising from increased vaccination coverage, and thus the reduction in hospitalisations and deaths.^7,8^

For the purposes of estimating hospitalisations and deaths, infection data generated by the model was translated into cases using a 2:1 ratio, a proportion chosen to reflect estimated infection/case ratios in Australia in 2020,^11^ in the absence of more recent Omicron-specific data. The relationship between case numbers, and hospitalisation and death numbers, were derived from case data using United Kingdom (UK) Omicron variant health burden datasets.^18,27^ UK data was used given the larger SARS-CoV-2 Omicron infection rate in the UK compared to Australia, and is appropriate given similar population demographics and health systems in Australia and the UK. We used the UK COVID-19 health burden datasets from 31^st^ December 2021 to 15^th^ January 2022 when (effectively) all SARS-CoV-2 transmission was due to the Omicron variant.^18^

The agent-based model represents the ages of each of the 273,407 persons in the modelled community (Newcastle, Australia) in 10 age bands, collected in the Australian census and accessed from the Australian Bureau of Statistics.^28,29^ Age-specific UK Omicron variant health burden data establish age-specific case/hospitalisation and case/mortality ratios, and these were used to assign age-specific health burden outcomes to each of the individuals infected within the model.^18^ Intensive Care Unit (ICU) demand was assumed to be one tenth of overall hospitalisation demand, a ratio derived from Australian data sources.^19^ These ratios are detailed in Table S1.

The estimated health burden outcome resulting from a range of alternative Western Australia border opening dates was determined by applying the simulation model software with a “burst” of infectious individuals introduced into the community after a specific opening date. Alternative opening date scenarios were evaluated, monthly from early February onwards. This permitted the effect which delaying border opening may have on the resulting health burden, and thus pressure on the health care system. These opening scenarios were simulated by seeding infectious individuals into the model to initiate a sustained COVID-19 Omicron outbreak in an effectively SARS-CoV-19 naive population. Daily infection data output from these simulation experiments were then used to estimate age-specific hospitalisation and mortality rates and ICU demand, for each of the border opening dates and are presented in the Results.

### ICU demand

The ability of health systems to successfully manage surges in cases and mitigate resulting morbidity and mortality rates due to highly transmissible SARS-CoV-19 variants is highly dependent on Intensive Care Unit (ICU) capacity, rather than general ward capacity. A survey conducted by the Australian and New Zealand Intensive Care Society from 30th August to 9^th^ September 2021 provides data on additional ICU bed and staff resources in Australian ICUs which permit capacity to increase in response to increased pandemic demand.^30^ Using ICU data from this survey we determined which Western Australia border opening strategies prevent ICU facilities from being overwhelmed following a surge in Omicron variant cases.

Western Australia’s maximum ICU bed capacity of 484 includes 159 current fully staffed beds, 52 additional physical ICU beds, with 273 further bed spaces in surge areas outside of ICUs.^30^ However, availability of medical and ICU nursing staff would only permit ∼31 additional ICU beds to open at pre-pandemic staffing levels.^30^ Staff resources may permit ICU facilities to exceed the 31 ICU bed threshold and open all 52 additional available ICU beds to meet excess demand, at least as a short-term response.

## Results

This study presents evidence on the impact on the health burden predicted to result from opening the borders of Western Australia at monthly intervals from 5^th^ February 2022 onwards, and the resulting rapid introduction of the SARS-CoV-2 Omicron variant into the state. Detailed estimates of the number of cases, hospitalisations, ICU admissions and deaths resulting from delaying border opening are presented in Table 2 and illustrated in Figure 3. These data indicate that opening the border later than early May 2022 will likely result in an increased health burden compared to earlier opening dates. For an early June opening date onwards, all health burden metrics are seen to increase steadily with time, see Figure 3. Prior to a May opening date, the health burden metrics remain steady with only slight increases over time. This corresponds to the plateau in estimated population-wide immunity which is reached once all second vaccine doses and 37.5% (3 out of 8 months of third dose rollout) of third doses have been administered to applicable age groups. This is predicted to occur by the end of June 2022, and before the waning effect from second and third doses begins to dominate, an immunity feature illustrated in Figure 1B.

**Table 2.** Health burden outcomes for alternative Western Australia border opening dates. All data scaled to Western Australia population of 2.7 million persons.

Cases
| Opening Date | Total | 0-12y | 13-24y | 25-44y | 45-64y | 65-79y | 80y+ |
| --- | --- | --- | --- | --- | --- | --- | --- |
| Feb 5 | 741,190 | 172,765 | 115,253 | 183,546 | 173,119 | 67,696 | 29,006 |
| Mar 5 | 754,774 | 173,989 | 117,782 | 187,075 | 176,920 | 69,622 | 29,804 |
| Apr 5 | 764,854 | 174,807 | 119,740 | 189,870 | 179,522 | 70,888 | 30,315 |
| May 5 | 806,634 | 177,988 | 127,518 | 201,033 | 191,421 | 76,624 | 32,897 |
| Jun 5 | 885,352 | 182,929 | 141,880 | 223,131 | 213,001 | 86,690 | 37,503 |
| Jul 5 | 961,278 | 186,884 | 156,217 | 244,292 | 235,223 | 97,154 | 41,941 |
| Aug 5 | 1,041,178 | 190,284 | 170,688 | 267,458 | 257,809 | 108,047 | 46,790 |
| Sep 5 | 1,107,195 | 192,638 | 183,289 | 286,555 | 276,649 | 117,252 | 50,914 |

Hospitalisations
| Opening Date | Total | 0-12y | 13-24y | 25-44y | 45-64y | 65-79y | 80y+ |
| --- | --- | --- | --- | --- | --- | --- | --- |
| Feb 5 | 2,861 | 87 | 123 | 343 | 507 | 730 | 1,071 |
| Mar 5 | 2,932 | 88 | 126 | 349 | 518 | 750 | 1,101 |
| Apr 5 | 2,980 | 88 | 128 | 354 | 526 | 764 | 1,120 |
| May 5 | 3,203 | 89 | 136 | 375 | 560 | 826 | 1,215 |
| Jun 5 | 3,603 | 92 | 152 | 416 | 624 | 934 | 1,385 |
| Jul 5 | 4,002 | 94 | 167 | 456 | 689 | 1,047 | 1,549 |
| Aug 5 | 4,425 | 95 | 183 | 499 | 755 | 1,165 | 1,728 |
| Sep 5 | 4,782 | 96 | 196 | 535 | 810 | 1,264 | 1,881 |

ICU Admissions
| Opening Date | Total | 0-12y | 13-24y | 25-44y | 45-64y | 65-79y | 80y+ |
| --- | --- | --- | --- | --- | --- | --- | --- |
| Feb 5 | 286 | 9 | 12 | 34 | 51 | 73 | 107 |
| Mar 5 | 293 | 9 | 13 | 35 | 52 | 75 | 110 |
| Apr 5 | 298 | 9 | 13 | 35 | 53 | 76 | 112 |
| May 5 | 320 | 9 | 14 | 38 | 56 | 83 | 122 |
| Jun 5 | 360 | 9 | 15 | 42 | 62 | 93 | 139 |
| Jul 5 | 400 | 9 | 17 | 46 | 69 | 105 | 155 |
| Aug 5 | 443 | 10 | 18 | 50 | 76 | 117 | 173 |
| Sep 5 | 478 | 10 | 20 | 54 | 81 | 126 | 188 |

Deaths
| Opening Date | Total | 0-12y | 13-24y | 25-44y | 45-64y | 65-79y | 80y+ |
| --- | --- | --- | --- | --- | --- | --- | --- |
| Feb 5 | 373 | 0 | 0 | 5 | 44 | 104 | 219 |
| Mar 5 | 383 | 0 | 0 | 5 | 45 | 107 | 225 |
| Apr 5 | 390 | 0 | 0 | 5 | 46 | 109 | 228 |
| May 5 | 421 | 0 | 1 | 6 | 49 | 118 | 248 |
| Jun 5 | 478 | 0 | 1 | 6 | 54 | 134 | 283 |
| Jul 5 | 534 | 0 | 1 | 7 | 60 | 150 | 316 |
| Aug 5 | 594 | 0 | 1 | 8 | 66 | 167 | 353 |
| Sep 5 | 644 | 0 | 1 | 8 | 71 | 181 | 384 |

Our model-generated data indicate that there is little benefit from delaying border opening, in terms of a reduction in the overall health burden, by one month from 5^th^ February 2022, the planned opening date, to the actual opening date of 3^rd^ March 2022. Results from our modelling analyses suggest that delaying border reopening even by one month may result in a slight increase in the health burden, see Table 2.

### Hospital and ICU demand

For a 5^th^ March 2022 border reopening, the peak in hospitalisations and ICU bed demand is predicted to occur approximately 2 months after Omicron variant cases are introduced into the naïve population. Health-care demand is estimated to peak at 510 general hospital beds and 51 ICU beds, Figures 3B and 3C respectively. Day by day predicted ICU bed demand for this opening date, along with the 10^th^ and 90^th^ percentiles, can be seen in Table 3.

**Table 3.** Daily ICU demand for 5^th^ March 2022 border opening

| Date | 10<sup>th</sup> Percentile | Median | 90<sup>th</sup> Percentile |
| --- | --- | --- | --- |
| 21/04/2022 | 2 | 8 | 20 |
| 22/04/2022 | 2 | 9 | 23 |
| 23/04/2022 | 3 | 10 | 27 |
| 24/04/2022 | 3 | 12 | 31 |
| 25/04/2022 | 4 | 15 | 35 |
| 26/04/2022 | 4 | 17 | 40 |
| 27/04/2022 | 5 | 20 | 43 |
| 28/04/2022 | 6 | 23 | 47 |
| 29/04/2022 | 8 | 27 | 51 |
| 30/04/2022 | 10 | 31 | 53 |
| 1/05/2022 | 11 | 35 | 55 |
| 2/05/2022 | 13 | 40 | 57 |
| 3/05/2022 | 16 | 44 | 57 |
| 4/05/2022 | 18 | 47 | 58 |
| 5/05/2022 | 22 | 49 | 58 |
| 6/05/2022 | 25 | 51 | 58 |
| 7/05/2022 | 27 | 51 | 58 |
| 8/05/2022 | 28 | 50 | 58 |
| 9/05/2022 | 28 | 50 | 58 |
| 10/05/2022 | 27 | 49 | 59 |
| 11/05/2022 | 24 | 47 | 58 |
| 12/05/2022 | 22 | 45 | 58 |
| 13/05/2022 | 20 | 43 | 57 |
| 14/05/2022 | 18 | 39 | 57 |
| 15/05/2022 | 16 | 35 | 56 |
| 16/05/2022 | 14 | 32 | 55 |
| 17/05/2022 | 11 | 29 | 54 |
| 18/05/2022 | 9 | 25 | 52 |
| 19/05/2022 | 8 | 22 | 50 |
| 20/05/2022 | 7 | 19 | 48 |
| 21/05/2022 | 6 | 16 | 44 |
| 22/05/2022 | 5 | 13 | 41 |
| 23/05/2022 | 4 | 11 | 37 |
| 24/05/2022 | 3 | 9 | 33 |
| 25/05/2022 | 3 | 8 | 29 |
| 26/05/2022 | 2 | 6 | 25 |
| 27/05/2022 | 2 | 5 | 21 |

Two key ICU demand thresholds for Western Australia are the 31 currently available (excess) fully staffed ICU beds, and the 52 physically available ICU beds which could be staffed by reassigned staff from other areas which can be maintained for approximately one month, as described in Litton, Huckson ^30^ These thresholds are highlighted by the yellow and orange horizontal lines in Figure 3C respectively. This daily ICU demand suggests that the 52 additional ICU beds available in Western Australia capable of being fully staffed in a surge situation will meet the maximum demand. This is due to the short duration of the ICU peak, with approximately 12 days over an arbitrary 40-bed requirement, and all below the median peak demand of 51 fully staffed ICU beds, as in Table 3. This table also presents the 90^th^ percentile confidence interval of the predicted ICU demand, indicating a low probability, worst-case outcome with a maximum daily demand at 59 ICU beds and 18 days over the surge capacity 52 bed threshold.

If the Western Australia border reopening was delayed until June 2022, expected hospitalisations would increase to ∼750, and ICU surge capacity would be exceeded, with ∼75 fully staffed ICU beds required. This increased pressure on the Western Australia health system would further increase for later border opening dates, reaching ∼1200 hospitalisations with a requirement of ∼120 ICU beds resulting from a September reopening. This is significantly higher than the ICU surge capacity of 52, which is restricted by availability of appropriately trained staff, a key constraint highlighted by Litton and his ICU colleagues.^30^

All health burden metrics follows a similar trajectory to that of ICU demand, with approximately constant numbers for February to May opening dates, and then a steady increase resulting from a June opening and later, as in Table 2. For a 5^th^ March 2022 opening a total of 2,932 hospitalisations, 293 ICU admissions and 383 deaths are estimated to occur, with similar numbers each month until a May reopening, Table 2. With a June 2022 opening date total numbers are estimated to increase to 3,603 hospitalisations, 360 ICU admissions and 478 deaths, which for a September opening date further increases to 4,782 hospitalisations, 478 ICU admissions and 644 deaths in a Western Australia population of ∼2.7 million persons. 80% confidence intervals for these values can be seen in Table S2; these confidence intervals are relatively small, less than 2% deviation from the median. Daily predicted ICU demand for a 5^th^ March 2022 border opening is presented in Table 3.

## Discussion

Western Australia was the only Australian state which maintaining a closed border to the rest of the Australia and to all other countries until early March 2022, resulted in limited SARS-CoV-2 transmission. This study conducted a comprehensive modelling analysis of the population-wide effects of alternative delayed border reopening dates. The aim was to determine an optimum reopening date, one which minimises the resulting Omicron variant health burden, and to understand whether the Western Australian hospital system would be equipped to handle the resulting peak in demand.

The use of an agent-based model permitted us to calculate the age-specific health burden under a range of border reopening dates. The ages of those infected, together with case/hospitalisation and case/fatality data for the Omicron variant,^18^ were used to determine the number of hospitalisations and fatalities, and demand for critical care in ICU facilities that may result.

### Managing the health burden

A key metric observed during an epidemic is the increased demand it makes on the health system, particularly the availability of additional general hospital and ICU beds, and suitably trained staff resources, to manage rapidly increasing demand. Following the Government of Western Australian opening interstate and international borders on 3^rd^ March 2022,^1^ the impact this opening date may have on the Western Australia hospital system is reflected using our modelled 5^th^ March opening date. Whilst there was prior ongoing local Omicron transmission in Western Australia prior to the opening date,^1^ a situation not directly modelled given its limited scale, the results are applicable to the emerging Omicron epidemic in Western Australia from late February 2022 onwards.

Our results predict that at the peak of the outbreak, 510 additional hospital beds will be required, 51 of which will be in ICUs. This is within the 52 additional physical ICU beds available^30^, but above the 31 beds that are currently able to be fully staffed. However, there is the ability to fully staff all 52 additional ICU beds using additional health system staff provided it is for a limited duration,^30^ Table 3. ICU demand following a 5^th^ March 2022 border opening in a worst-case scenario is shown by the 90^th^ percentile confidence interval column in Table 3, indicating a maximum daily demand of 59 ICU beds, with 18 days over the 52-bed surge threshold. These data suggests that Western Australia will have adequate ICU capacity to meet this forecast demand, given: *1)* the low probability of reaching this 59 bed demand peak, *2)* the limited duration above 52 beds, and *3)* current availability of greater than 52 physical available ICU beds which may be used if additional staff can be found. As discussed by Litton and his Australian ICU colleagues, it is the availability of specialist ICU staff which is the limiting factor, rather than physical ICU beds.^30^ The limited duration of this worst-case peak demand makes it highly likely that additional staff resources will be available for this short-duration, high ICU demand period.

This short-term surge in excess ICU demand is likely to be met with available staff resources, however it may be impacted by the requirement for COVID positive and close contact ICU staff to isolate. As has been seen in previous Omicron epidemics in Victoria, New South Wales, Queensland and South Australia,^31^ the need for health care workers to isolate may have a significant impact on the ability of Western Australia to respond to the peak of the Omicron outbreak.

We have applied a relatively conservative hospitalisation/ICU ratio of 10:1 to estimate peak ICU demand. In reality, this number fluctuates depending on the state (or country) and the date when it is measured.^19,32^ This variation is likely due to different vaccination rates, age demographics, the proportion of Delta to Omicron infections, and new treatment options such as use of antiviral drug therapy.^33^ In general, the Omicron hospitalisation/ICU ratio has been observed to start higher in early January 2022, then trended downwards before reaching a plateau at the beginning of February 2022, likely due to the changing factors listed above.^19,32^ Thus 10:1 may be a somewhat pessimistic estimate, and an 8:1 ratio may be closer to reality, 20% lower than used in our ICU demand analyses.

Comparing Queensland and South Australian fatality rates against the predictions for Western Australia (Figure 2D) the peak in actual Omicron implicated daily deaths is something of an overestimation. This suggests that the earlier UK Omicron case/fatality data used overestimates the fatality rate by 20-40%.^18^ This indicates that COVID-19 treatment options, such as recently introduced antiviral drug therapy, may have reduced the previous Omicron fatality rate.^34^

It is well understood that protection against symptomatic disease from vaccination for the Omicron variant is lower than for Delta, and declines at a greater rate.^4^ Results from our study illustrate that without additional immunity afforded from natural infection, population-wide immunity will decrease rapidly approximately four months after a third dose booster program has been completed, as in Figure 1. This may be assumed to result in an increase in cases, and thus attendant health burden metrics. While we have assumed constant age-specific case/hospitalisation and case/fatality ratios, in reality protection against these outcomes will wane at different rates compared to symptomatic disease, typically more slowly. Emerging data from the UK supports this,^35^ and supports the qualitative result that health burdens will increase over time in an initially vaccinated but COVID-19 naive population.

Omicron is generally understood to have a transmission advantage but is less pathogenic than Delta,^16,17,36,37^ however quantifying and thus comparing these parameters is difficult given differences in the populations in which they are circulating, and availability and access to early characterising data. While Omicron infections are dominating Delta infections in most parts of the world, its reproduction advantage may be due to increased inherent transmissibility, increased immunity escape, a different population response to arrival of Omicron, or some combination of these.^37^ To manage this uncertainty, we employed the calibration process described in the methods section. This exploited the availability of detailed Omicron epidemic data from two Australian states, Queensland and South Australia. These states opened their borders earlier than Western Australia, but after the Omicron variant had already established itself in Australia’s two largest cities, Sydney (New South Wales) and Melbourne (Victoria).^24,25^ This approach allowed us to take into account self-adopted population behaviour changes, such as working-from-home and lessened contact in the community, as captured by Google mobility data.^26^ This calibration process utilised definitive Omicron variant hospitalisation data from South Australia and Queensland and observed vaccination rates, to obtain an effective reproduction number R_eff_ which excluded vaccination-induced immunity. This derived R_eff_ was applied with Western Australia vaccination data to predict daily Omicron infections, and from these data to estimate the resulting health burden.

### Delta and Omicron comparison

A previous study by the authors evaluated the health burden of the Delta variant under alternative vaccination coverage levels, with and without activation of non-pharmaceutical interventions (NPIs).^8^ While that study did not model the effects of waning immunity following vaccination, which suggest that its results are likely to be somewhat optimistic, they can be compared none the less. That Delta study estimated a peak of ∼9,000 cases a day in a population the size of Western Australia (∼2.7 million persons) without NPIs activated. This compares to a ∼30,000 peak forecast for the Omicron variant, assuming a low level of NPIs in effect. Despite the three-fold difference in peak daily cases, hospital bed demand in Western Australia is roughly comparable, with a peak of ∼540 beds required for a Delta outbreak, compared to ∼510 for an Omicron outbreak. A major difference lies in expected ICU bed demand, with the hospitalisation/ICU ratio for Delta infections assumed to be 5:1, meaning a peak of 540 hospital beds would include 108 ICU beds, double the number available at Western Australia’s surge capacity of 52.^30^ With a higher case/fatality ratio, the total number of deaths estimated in a Delta epidemic is ∼230 in a population the size of Western Australia, compared to ∼380 in this Omicron study. This is a significant difference, however this difference may be explained by three factors: the significantly higher infection rate and case numbers due to Omicron’s greater transmissibility; reduced vaccine effectiveness compared to Delta; and the absence of waning immunity in our prior Delta variant study, whose inclusion is likely to increase the Delta variant health burden.^8^

### Policy implications

Our results highlight the benefit of reopening the Western Australia borders earlier rather than later. This is due to the optimal level of vaccine-induced immunity within the population, a function of the State’s very high third dose vaccination rate, and before immunity gained from vaccination begins to wane. Our results also highlight the necessity of replacing immunity derived from vaccination by immunity resulting from SARS-CoV-2 Omicron variant infection. That is, for those whose immunity has waned, and they become susceptible to infection once more, their immunity is boosted following infection. It is also hoped that naturally derived immunity will provide better protection against both reinfection and symptomatic illness compared to that derived from vaccination, thus giving longer lasting protection. Data gathered over the next 12 months is needed to better understand such protective effects.

The announcement by the Government of Western Australia of a 3^rd^ March 2022 border opening date^1^ was a welcome decision, given our findings. Additionally, this decision was made in the context of an ongoing local Omicron variant outbreak which started to grow exponentially prior to the opening date.^1^ Whilst our modelling does not handle this specific scenario, our results generally apply to a population where an outbreak occurs with a given population immunity whether that occurs as a result of a hard border being removed or “leakage” through an existing hard border. Given the plateau in population immunity determined (Figure 1B) and the resulting epidemic curves (Figure 3) these results may be used to predict the increased health burden arising from Western Australia’s Omicron epidemic, and plan health system responses. We further expect that the predicted daily ICU demand (Table 3) will be indicative of the daily ICU load during the peak of the current Western Australian outbreak. The earlier peak in the Western Australia Omicron epidemic resulting from an early border reopening has an additional benefit; it allows the predicted Omicron outbreak peak to occur before any potential influenza epidemic, which generally occurs from May onwards during the Southern Hemisphere winter months. This should prevent the simultaneous growth of COVID-19 and influenza case numbers, thus lessening pressure on the health system and available hospital resources. Had the borders opened in early February as originally planned, this would also have been within the optimal window for border reopening.

### Limitations

This study utilised Omicron data publicly available from mid-January 2022 up to late February 2022. Given that the emergence of this variant occurred in November 2021, further data may allow us to refine our findings. For example, there is limited data on the duration of immunity resulting from Omicron infection, and the long-term waning effects of booster vaccinations.

In the absence of further data, we assumed protection against symptomatic disease to be a proxy for vaccine effectiveness against infection. We have used UK data sources given their rapid availability, and the scale and earlier occurrence of Omicron outbreaks in the UK compared to Australia. Use of UK data is appropriate given the similar population demographics, healthcare systems, and vaccination rates in both countries.

We have assumed that the behaviour of the population does not change over the course of the modelled epidemic, and that limited mandated physical distancing measures and self-adopted social distancing remains constant. This level of NPI measures is implicitly modelled using data from Omicron outbreaks in two Australian states to derive our effective reproduction number, and thus the probability of transmission between infectious/susceptible pairs of individuals. However, this assumes population behaviour remains constant over the course of an outbreak, which may not be the case. Awareness of increasing case numbers may cause individuals to reduce movement and contacts and lead to “shadow lockdowns”, while decreasing case numbers may lead to increased movement and mixing in public spaces.^38^

## Conclusions

This study determined the impact of delaying the Western Australian border reopening in terms of the resulting health burden, in particular peak ICU bed demand. Modelling the extremely high Western Australian vaccination rate and documented vaccine-induced immunity waning rates, we determined a period where the overall population immunity is at its highest, between February 2022 and June 2022. Border opening occurring prior to this period ending resulted in the lowest health burden in comparison to opening in June 2022 and later. Opening the border on 5^th^ March 2022 was predicted to result in peak ICU demand of 51 beds, just below the available surge capacity in Western Australian ICUs. On 18th February 2022 the Government of Western Australia announced the borders would be reopened on 3^rd^ March 2022 amid an ongoing local outbreak, and results presented for a 5^th^ March 2022 opening predict the likely resulting health burden in the State. This study further highlights the challenge of relying solely on immunity derived from vaccination, and the rapid waning of such immunity at the individual level, without “compensating” immunity from infection. This suggests that for future SARS-CoV-2 variant epidemics, such as those which are both highly transmissible and highly pathogenic, immunity derived from infection is necessary. The resulting health burden will then need to be managed by applying physical distancing NPI measures, to lower the peak in hospitalisations, and by treatment with antiviral drugs, to reduce the severity of illness and thus ICU demand. Such treatment options are important to protect the elderly and other at-risk groups, those who have a poorer immune response to vaccination. The rapid detection of infections in the elderly, such as those in Residential Aged Care Facilities, and then timely treatment, is required.

## Data Availability

All data referred to is publicly available.

## Supplementary Data

### Case Hospitalisation/Fatality Ratios

**Table S1.** Age specific Case/Hospitalisation and Case/Fatality ratios as derived from UK age specific case, hospitalisation and fatality data.^18,27^

| Age group | Case / Hospitalisation Ratio | Case / Fatality Ratio |
| --- | --- | --- |
| 0 – 6 months | $0.000 \times 10^{-4}$ | $0.000 \times 10^{-4}$ |
| 7 – 24 months | $0.000 \times 10^{-4}$ | $0.000 \times 10^{-4}$ |
| 3 – 5 years | $1.221 \times 10^{-3}$ | $4.770 \times 10^{-7}$ |
| 6 – 12 years | $3.673 \times 10^{-4}$ | $4.870 \times 10^{-7}$ |
| 13 – 17 years | $1.003 \times 10^{-3}$ | $3.523 \times 10^{-6}$ |
| 18 – 24 years | $1.115 \times 10^{-3}$ | $4.489 \times 10^{-6}$ |
| 25 – 44 years | $1.866 \times 10^{-3}$ | $2.893 \times 10^{-5}$ |
| 45 – 64 years | $2.928 \times 10^{-3}$ | $2.558 \times 10^{-4}$ |
| 65 – 79 years | $1.078 \times 10^{-2}$ | $1.541 \times 10^{-3}$ |
| 80+ years | $3.694 \times 10^{-2}$ | $7.535 \times 10^{-3}$ |

**Table S2:** Health burden outcomes for alternative Western Australia border opening dates. All numbers are scaled to a WA population of 2.7 million, and the 10^th^ and 90^th^ percentiles are included in brackets.

| Cases |  |  |  |  |  |  |  |
| --- | --- | --- | --- | --- | --- | --- | --- |
| Opening |  |  |  |  |  |  |  |
| Date | Total | 0-12y | 13-24y | 25-44y | 45-64y | 65-79y | 80y+ |
| Feb 5 | 741,190<br>[736,184-747,401] | 172,765<br>[171,750-173,811] | 115,253<br>[113,849-116,773] | 183,546<br>[181,876-185,309] | 173,119<br>[171,453-174,722] | 67,696<br>[66,750-68,834] | 29,006<br>[28,549-29,547] |
| Mar 5 | 754,774<br>[750,956-759,073] | 173,989<br>[173,091-174,870] | 117,782<br>[116,530-119,151] | 187,075<br>[185,868-188,455] | 176,920<br>[175,606-178,156] | 69,622<br>[68,813-70,539] | 29,804<br>[29,254-30,290] |
| Apr 5 | 764,854<br>[761,361-1,108,974] | 174,807<br>[173,802-191,927] | 119,740<br>[118,488-184,353] | 189,870<br>[188,722-290,013] | 179,522<br>[178,399-280,442] | 70,888<br>[69,962-113,349] | 30,315<br>[29,895-48,639] |
| May 5 | 806,634<br>[786,518-1,128,621] | 177,988<br>[176,302-192,703] | 127,518<br>[123,423-188,537] | 201,033<br>[195,221-295,858] | 191,421<br>[185,209-285,736] | 76,624<br>[73,828-116,266] | 32,897<br>[31,598-50,105] |
| Jun 5 | 885,352<br>[867,659-906,399] | 182,929<br>[181,625-184,329] | 141,880<br>[138,468-145,707] | 223,131<br>[217,773-229,043] | 213,001<br>[208,416-219,138] | 86,690<br>[84,425-89,929] | 37,503<br>[36,385-38,858] |
| Jul 5 | 961,278<br>[945,719-982,769] | 186,884<br>[185,924-187,996] | 156,217<br>[152,828-159,952] | 244,292<br>[239,849-250,832] | 235,223<br>[231,001-240,988] | 97,154<br>[94,765-99,880] | 41,941<br>[40,880-43,231] |
| Aug 5 | 1,041,178<br>[1,027,168-1,058,926] | 190,284<br>[189,532-191,168] | 170,688<br>[168,209-174,447] | 267,458<br>[263,080-272,643] | 257,809<br>[253,679-262,681] | 108,047<br>[106,050-110,764] | 46,790<br>[45,820-47,875] |
| Sep 5 | 1,107,195<br>[1,096,916-1,127,152] | 192,638<br>[192,120-193,377] | 183,289<br>[181,072-187,018] | 286,555<br>[283,256-292,130] | 276,649<br>[273,620-282,977] | 117,252<br>[115,905-120,267] | 50,914<br>[50,210-52,200] |

Hospitalisations
| Opening Date | Total | 0-12y | 13-24y | 25-44y | 45-64y | 65-79y | 80y+ |
| --- | --- | --- | --- | --- | --- | --- | --- |
| Feb 5 | 2,861<br>[2,824-2,903] | 87<br>[86-87] | 123<br>[122-125] | 343<br>[339-346] | 507<br>[502-512] | 730<br>[719-742] | 1,071<br>[1,055-1,091] |
| Mar 5 | 2,932<br>[2,895-2,968] | 88<br>[87-88] | 126<br>[125-128] | 349<br>[347-352] | 518<br>[514-522] | 750<br>[742-760] | 1,101<br>[1,081-1,119] |
| Apr 5 | 2,980<br>[2,947-4,674] | 88<br>[87-96] | 128<br>[127-197] | 354<br>[352-541] | 526<br>[522-821] | 764<br>[754-1,222] | 1,120<br>[1,104-1,797] |
| May 5 | 3,203<br>[3,090-4,791] | 89<br>[89-97] | 136<br>[132-202] | 375<br>[364-552] | 560<br>[542-837] | 826<br>[796-1,253] | 1,215<br>[1,167-1,851] |
| Jun 5 | 3,603<br>[3,510-3,722] | 92<br>[91-92] | 152<br>[148-156] | 416<br>[406-427] | 624<br>[610-642] | 934<br>[910-969] | 1,385<br>[1,344-1,435] |
| Jul 5 | 4,002<br>[3,912-4,112] | 94<br>[93-94] | 167<br>[164-171] | 456<br>[448-468] | 689<br>[676-706] | 1,047<br>[1,021-1,077] | 1,549<br>[1,510-1,597] |
| Aug 5 | 4,425<br>[4,344-4,522] | 95<br>[95-96] | 183<br>[180-187] | 499<br>[491-509] | 755<br>[743-769] | 1,165<br>[1,143-1,194] | 1,728<br>[1,693-1,768] |
| Sep 5 | 4,782<br>[4,724-4,895] | 96<br>[96-97] | 196<br>[194-200] | 535<br>[529-545] | 810<br>[801-828] | 1,264<br>[1,249-1,296] | 1,881<br>[1,855-1,928] |

ICU Admissions
| Opening Date | Total | 0-12y | 13-24y | 25-44y | 45-64y | 65-79y | 80y+ |
| --- | --- | --- | --- | --- | --- | --- | --- |
| Feb 5 | 286<br>[282-290] | 9<br>[9-9] | 12<br>[12-13] | 34<br>[34-35] | 51<br>[50-51] | 73<br>[72-74] | 107<br>[105-109] |
| Mar 5 | 293<br>[290-297] | 9<br>[9-9] | 13<br>[12-13] | 35<br>[35-35] | 52<br>[51-52] | 75<br>[74-76] | 110<br>[108-112] |
| Apr 5 | 298<br>[295-467] | 9<br>[9-10] | 13<br>[13-20] | 35<br>[35-54] | 53<br>[52-82] | 76<br>[75-122] | 112<br>[110-180] |
| May 5 | 320<br>[309-479] | 9<br>[9-10] | 14<br>[13-20] | 38<br>[36-55] | 56<br>[54-84] | 83<br>[80-125] | 122<br>[117-185] |
| Jun 5 | 360<br>[351-372] | 9<br>[9-9] | 15<br>[15-16] | 42<br>[41-43] | 62<br>[61-64] | 93<br>[91-97] | 139<br>[134-144] |
| Jul 5 | 400<br>[391-411] | 9<br>[9-9] | 17<br>[16-17] | 46<br>[45-47] | 69<br>[68-71] | 105<br>[102-108] | 155<br>[151-160] |
| Aug 5 | 442<br>[434-452] | 10<br>[9-10] | 18<br>[18-19] | 50<br>[49-51] | 75<br>[74-77] | 116<br>[114-119] | 173<br>[169-177] |
| Sep 5 | 478<br>[472-489] | 10<br>[10-10] | 20<br>[19-20] | 53<br>[53-55] | 81<br>[80-83] | 126<br>[125-130] | 188<br>[185-193] |

Deaths
| Opening Date | Total | 0-12y | 13-24y | 25-44y | 45-64y | 65-79y | 80y+ |
| --- | --- | --- | --- | --- | --- | --- | --- |
| Feb 5 | 373<br>[368-379] | 0<br>[0-0] | 0<br>[0-0] | 5<br>[5-5] | 44<br>[44-45] | 104<br>[103-106] | 219<br>[215-223] |
| Mar 5 | 383<br>[377-389] | 0<br>[0-0] | 0<br>[0-0] | 5<br>[5-5] | 45<br>[45-46] | 107<br>[106-109] | 225<br>[220-228] |
| Apr 5 | 390<br>[385-622] | 0<br>[0-0] | 0<br>[0-1] | 5<br>[5-8] | 46<br>[46-72] | 109<br>[108-175] | 228<br>[225-366] |
| May 5 | 421<br>[405-639] | 0<br>[0-0] | 1<br>[1-1] | 6<br>[6-9] | 49<br>[47-73] | 118<br>[114-179] | 248<br>[238-378] |
| Jun 5 | 478<br>[465-495] | 0<br>[0-0] | 1<br>[1-1] | 6<br>[6-7] | 54<br>[53-56] | 134<br>[130-139] | 283<br>[274-293] |
| Jul 5 | 534<br>[521-549] | 0<br>[0-0] | 1<br>[1-1] | 7<br>[7-7] | 60<br>[59-62] | 150<br>[146-154] | 316<br>[308-326] |
| Aug 5 | 594<br>[582-607] | 0<br>[0-0] | 1<br>[1-1] | 8<br>[8-8] | 66<br>[65-67] | 167<br>[163-171] | 353<br>[345-361] |
| Sep 5 | 644<br>[636-660] | 0<br>[0-0] | 1<br>[1-1] | 8<br>[8-8] | 71<br>[70-72] | 181<br>[179-185] | 384<br>[378-393] |

### Worst case scenario new variant

To model a potential worst case scenario with Omicron’s transmissibility and Delta’s virulence we also applied to the same infection data utilised in the results section to hospitalisations, ICU and fatality ratios used in a previous paper.^8^ These data are presented below in Table S3, and the hospitalisation/ICU ratio was 5:1, with an average hospital stay of 8 days as described in Abdullah, Myers ^16^ and Maslo, Friedland ^17^.

**Table S3.** Age specific Case/Hospitalisation and Case/Fatality ratios for the Delta variant as previously derived from UK age specific case, hospitalisation and fatality data^8^

| Age group | Case / Hospitalisation Ratio | Case / Fatality Ratio |
| --- | --- | --- |
| 0 – 6 months | $0.0 \times 10^{-4}$ | $0.0 \times 10^{-6}$ |
| 7 – 24 months | $0.0 \times 10^{-4}$ | $0.0 \times 10^{-6}$ |
| 3 – 5 years | $1.956 \times 10^{-3}$ | $4.240 \times 10^{-6}$ |
| 6 – 12 years | $9.110 \times 10^{-4}$ | $3.704 \times 10^{-6}$ |
| 13 – 17 years | $1.501 \times 10^{-3}$ | $2.684 \times 10^{-5}$ |
| 18 – 24 years | $1.594 \times 10^{-3}$ | $3.976 \times 10^{-5}$ |
| 25 – 44 years | $6.415 \times 10^{-3}$ | $9.577 \times 10^{-5}$ |
| 45 – 64 years | $9.704 \times 10^{-3}$ | $9.660 \times 10^{-4}$ |
| 65 – 79 years | $2.031 \times 10^{-2}$ | $4.356 \times 10^{-3}$ |
| 80+ years | $5.192 \times 10^{-2}$ | $1.590 \times 10^{-2}$ |

**Figure S1:**
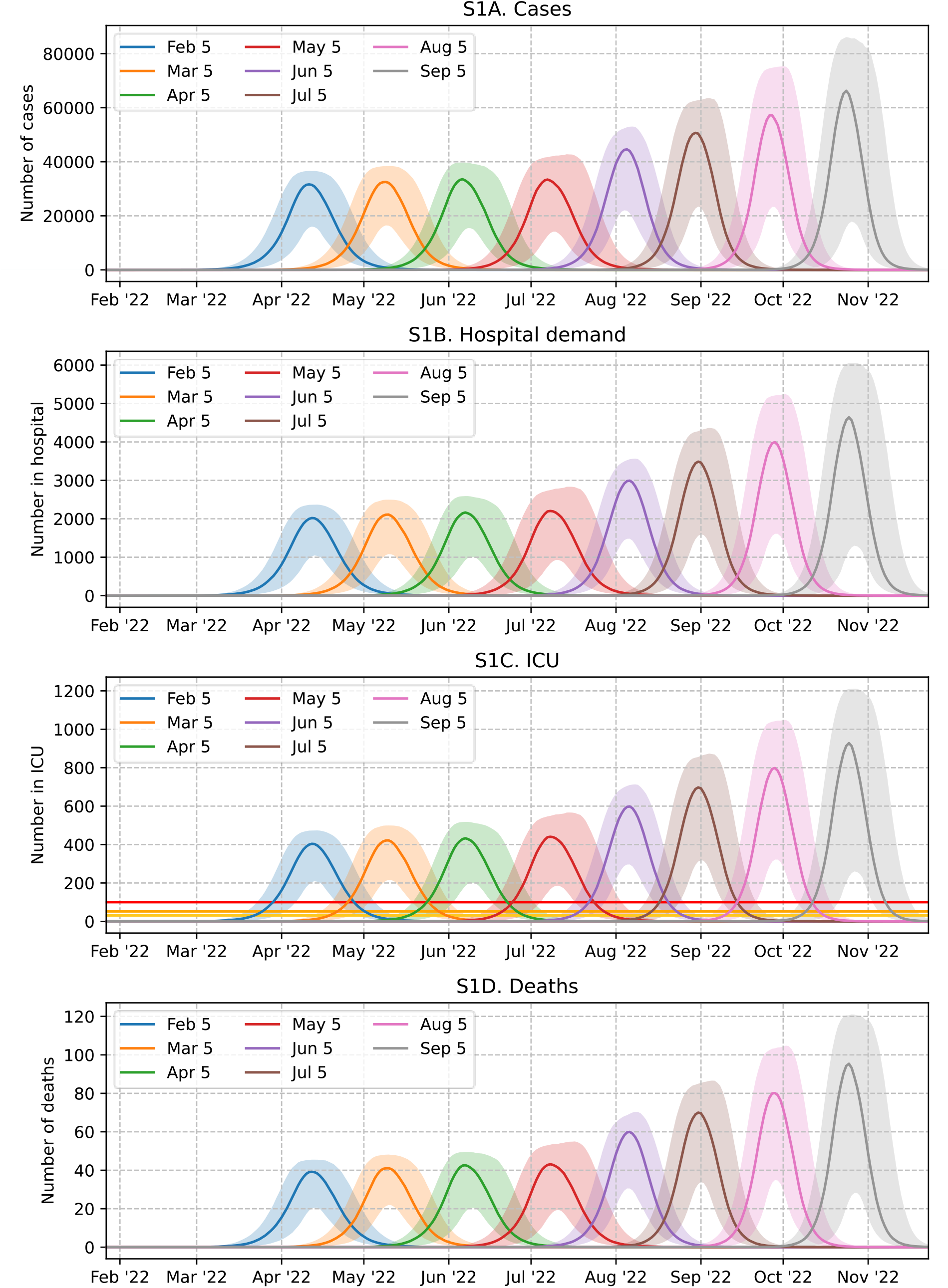
Predicated daily cases, hospitalisations, ICU demand and fatalities for Omicron epidemic in WA with alternative border opening dates using Delta variant hospitalisation and fatality rates. Vaccination at WA’s reported vaccination rate, with second doses administered over 8 months.

**Table S4:** Health burden outcomes for alternative Western Australia border opening dates with Delta hospitalisation, ICU and fatality rates. All numbers are scaled to a WA population of 2.7 million, and the 10^th^ and 90^th^ percentiles are included in brackets.

| **Cases** |  |  |  |  |  |  |  |
| --- | --- | --- | --- | --- | --- | --- | --- |
| **Opening Date** | **Total** | **0-12y** | **13-24y** | **25-44y** | **45-64y** | **65-79y** | **80y+** |
| Feb 5 | 741,190<br>[736,184-747,401] | 172,765<br>[171,750-173,811] | 115,253<br>[113,849-116,773] | 183,546<br>[181,876-185,309] | 173,119<br>[171,453-174,722] | 67,696<br>[66,750-68,834] | 29,006<br>[28,549-29,547] |
| Mar 5 | 754,774<br>[750,956-759,073] | 173,989<br>[173,091-174,870] | 117,782<br>[116,530-119,151] | 187,075<br>[185,868-188,455] | 176,920<br>[175,606-178,156] | 69,622<br>[68,813-70,539] | 29,804<br>[29,254-30,290] |
| Apr 5 | 764,854<br>[761,361-1,108,974] | 174,807<br>[173,802-191,927] | 119,740<br>[118,488-184,353] | 189,870<br>[188,722-290,013] | 179,522<br>[178,399-280,442] | 70,888<br>[69,962-113,349] | 30,315<br>[29,895-48,639] |
| May 5 | 806,634<br>[786,518-1,128,621] | 177,988<br>[176,302-192,703] | 127,518<br>[123,423-188,537] | 201,033<br>[195,221-295,858] | 191,421<br>[185,209-285,736] | 76,624<br>[73,828-116,266] | 32,897<br>[31,598-50,105] |
| Jun 5 | 885,352<br>[867,659-906,399] | 182,929<br>[181,625-184,329] | 141,880<br>[138,468-145,707] | 223,131<br>[217,773-229,043] | 213,001<br>[208,416-219,138] | 86,690<br>[84,425-89,929] | 37,503<br>[36,385-38,858] |
| Jul 5 | 961,278<br>[945,719-982,769] | 186,884<br>[185,924-187,996] | 156,217<br>[152,828-159,952] | 244,292<br>[239,849-250,832] | 235,223<br>[231,001-240,988] | 97,154<br>[94,765-99,880] | 41,941<br>[40,880-43,231] |
| Aug 5 | 1,041,178<br>[1,027,168-1,058,926] | 190,284<br>[189,532-191,168] | 170,688<br>[168,209-174,447] | 267,458<br>[263,080-272,643] | 257,809<br>[253,679-262,681] | 108,047<br>[106,050-110,764] | 46,790<br>[45,820-47,875] |
| Sep 5 | 1,107,195<br>[1,096,916-1,127,152] | 192,638<br>[192,120-193,377] | 183,289<br>[181,072-187,018] | 286,555<br>[283,256-292,130] | 276,649<br>[273,620-282,977] | 117,252<br>[115,905-120,267] | 50,914<br>[50,210-52,200] |

Hospitalisations
| **Opening Date** | **Total** | **0-12y** | **13-24y** | **25-44y** | **45-64y** | **65-79y** | **80y+** |
| --- | --- | --- | --- | --- | --- | --- | --- |
| Feb 5 | 6,085<br>[6,012-6,166] | 167<br>[166-168] | 179<br>[177-182] | 1,177<br>[1,167-1,189] | 1,680<br>[1,664-1,696] | 1,375<br>[1,356-1,398] | 1,506<br>[1,482-1,534] |
| Mar 5 | 6,230<br>[6,162-6,298] | 168<br>[167-169] | 183<br>[181-186] | 1,200<br>[1,192-1,209] | 1,717<br>[1,704-1,729] | 1,414<br>[1,398-1,433] | 1,548<br>[1,519-1,573] |
| Apr 5 | 6,329<br>[6,268-9,882] | 169<br>[168-185] | 186<br>[184-287] | 1,218<br>[1,211-1,860] | 1,742<br>[1,731-2,721] | 1,440<br>[1,421-2,303] | 1,574<br>[1,552-2,525] |
| May 5 | 6,782<br>[6,552-10,114] | 172<br>[170-186] | 199<br>[192-294] | 1,290<br>[1,252-1,898] | 1,858<br>[1,797-2,773] | 1,557<br>[1,500-2,362] | 1,708<br>[1,641-2,602] |
| Jun 5 | 7,604<br>[7,415-7,845] | 177<br>[175-178] | 221<br>[216-227] | 1,431<br>[1,397-1,469] | 2,067<br>[2,023-2,127] | 1,761<br>[1,715-1,827] | 1,947<br>[1,889-2,018] |
| Jul 5 | 8,425<br>[8,245-8,652] | 180<br>[179-181] | 243<br>[238-249] | 1,567<br>[1,539-1,609] | 2,283<br>[2,242-2,339] | 1,974<br>[1,925-2,029] | 2,178<br>[2,123-2,245] |
| Aug 5 | 9,291<br>[9,128-9,490] | 184<br>[183-184] | 266<br>[262-271] | 1,716<br>[1,688-1,749] | 2,502<br>[2,462-2,549] | 2,195<br>[2,154-2,250] | 2,429<br>[2,379-2,486] |
| Sep 5 | 10,020<br>[9,901-10,251] | 186<br>[185-187] | 285<br>[282-291] | 1,838<br>[1,817-1,874] | 2,685<br>[2,655-2,746] | 2,382<br>[2,355-2,443] | 2,644<br>[2,607-2,710] |

ICU Admissions
| Opening Date | Total | 0-12y | 13-24y | 25-44y | 45-64y | 65-79y | 80y+ |
| --- | --- | --- | --- | --- | --- | --- | --- |
| Feb 5 | 1,217<br>[1,202-1,233] | 33<br>[33-34] | 36<br>[35-36] | 235<br>[233-238] | 336<br>[333-339] | 275<br>[271-280] | 301<br>[296-307] |
| Mar 5 | 1,246<br>[1,232-1,260] | 34<br>[33-34] | 37<br>[36-37] | 240<br>[238-242] | 343<br>[341-346] | 283<br>[280-287] | 310<br>[304-315] |
| Apr 5 | 1,266<br>[1,254-1,976] | 34<br>[34-37] | 37<br>[37-57] | 244<br>[242-372] | 348<br>[346-544] | 288<br>[284-461] | 315<br>[310-505] |
| May 5 | 1,356<br>[1,310-2,023] | 34<br>[34-37] | 40<br>[38-59] | 258<br>[250-380] | 372<br>[359-555] | 311<br>[300-472] | 342<br>[328-520] |
| Jun 5 | 1,521<br>[1,483-1,569] | 35<br>[35-36] | 44<br>[43-45] | 286<br>[279-294] | 413<br>[405-425] | 352<br>[343-365] | 389<br>[378-404] |
| Jul 5 | 1,685<br>[1,649-1,730] | 36<br>[36-36] | 49<br>[48-50] | 313<br>[308-322] | 457<br>[448-468] | 395<br>[385-406] | 436<br>[425-449] |
| Aug 5 | 1,858<br>[1,826-1,898] | 37<br>[37-37] | 53<br>[52-54] | 343<br>[338-350] | 500<br>[492-510] | 439<br>[431-450] | 486<br>[476-497] |
| Sep 5 | 2,004<br>[1,980-2,050] | 37<br>[37-37] | 57<br>[56-58] | 368<br>[363-375] | 537<br>[531-549] | 476<br>[471-489] | 529<br>[521-542] |

Deaths
| Opening Date | Total | 0-12y | 13-24y | 25-44y | 45-64y | 65-79y | 80y+ |
| --- | --- | --- | --- | --- | --- | --- | --- |
| Feb 5 | 945<br>[932-961] | 1<br>[1-1] | 4<br>[4-4] | 18<br>[17-18] | 167<br>[166-169] | 295<br>[291-300] | 461<br>[454-470] |
| Mar 5 | 971<br>[957-984] | 1<br>[1-1] | 4<br>[4-4] | 18<br>[18-18] | 171<br>[170-172] | 303<br>[300-307] | 474<br>[465-482] |
| Apr 5 | 987<br>[975-1,573] | 1<br>[1-1] | 4<br>[4-6] | 18<br>[18-28] | 173<br>[172-271] | 309<br>[305-494] | 482<br>[475-773] |
| May 5 | 1,066<br>[1,026-1,614] | 1<br>[1-1] | 4<br>[4-7] | 19<br>[19-28] | 185<br>[179-276] | 334<br>[322-506] | 523<br>[502-797] |
| Jun 5 | 1,206<br>[1,174-1,249] | 1<br>[1-1] | 5<br>[5-5] | 21<br>[21-22] | 206<br>[201-212] | 378<br>[368-392] | 596<br>[578-618] |
| Jul 5 | 1,347<br>[1,315-1,385] | 1<br>[1-1] | 5<br>[5-6] | 23<br>[23-24] | 227<br>[223-233] | 423<br>[413-435] | 667<br>[650-687] |
| Aug 5 | 1,496<br>[1,467-1,530] | 1<br>[1-1] | 6<br>[6-6] | 26<br>[25-26] | 249<br>[245-254] | 471<br>[462-482] | 744<br>[728-761] |
| Sep 5 | 1,622<br>[1,601-1,662] | 1<br>[1-1] | 6<br>[6-6] | 27<br>[27-28] | 267<br>[264-273] | 511<br>[505-524] | 809<br>[798-830] |

## References

1. Carmody J, Weber D. WA border opening date is March 3, as Mark McGowan announces new COVID restrictions. 2022. https://www.abc.net.au/news/2022-02-18/mark-mcgowan-announces-wa-border-update/100843126 (accessed 21st February 2022).

2. Campbell F, Archer B, Laurenson-Schafer H, et al. Increased transmissibility and global spread of SARS-CoV-2 variants of concern as at June 2021. Eurosurveillance 2021; 26(24): 2100509.

3. Update on Omicron. 2021. https://www.who.int/news/item/28-11-2021-update-on-omicron (accessed 4th Feb 2022).

4. SARS-CoV-2 variants of concern and variants under investigation in England, Technical Briefing 34. In: Agency UHS, editor.; 2022. p. 22.

5. Li Q, Guan X, Wu P, et al. Early Transmission Dynamics in Wuhan, China, of Novel Coronavirus–Infected Pneumonia. New England Journal of Medicine 2020; 382(13): 1199–207.

6. Milne GJ, Xie S, Poklepovich D. A Modelling Analysis of Strategies for Relaxing COVID-19 Social Distancing. medRxiv 2020: 2020.05.19.20107425.

7. Milne GJ, Xie S, Poklepovich D, O’Halloran D, Yap M, Whyatt D. A modelling analysis of the effectiveness of second wave COVID-19 response strategies in Australia. Scientific Reports 2021; 11(1): 11958.

8. Milne GJ, Carrivick J, Whyatt D. Mitigating the SARS-CoV-2 Delta disease burden in Australia by non-pharmaceutical interventions and vaccinating children: a modelling analysis. BMC Med 2022; 20(1): 80.

9. Milne GJ, Kelso JK, Kelly HA, Huband ST, McVernon J. A small community model for the transmission of infectious diseases: comparison of school closure as an intervention in individual-based models of an influenza pandemic. PLoS One 2008; 3(12): e4005.

10. Anderson RM, May RM. Infectious diseases of humans: dynamics and control: Oxford university press; 1992.

11. Phipps SJ, Grafton RQ, Kompas T. Robust estimates of the true (population) infection rate for COVID-19: a backcasting approach. R Soc Open Sci 2020; 7(11): 200909.

12. Jansen L, Tegomoh B, Lange K, et al. Investigation of a SARS-CoV-2 B.1.1.529 (Omicron) Variant Cluster - Nebraska, November-December 2021. MMWR Morb Mortal Wkly Rep 2021; 70(5152): 1782–4.

13. Sah P, Fitzpatrick MC, Zimmer CF, et al. Asymptomatic SARS-CoV-2 infection: A systematic review and meta-analysis. Proceedings of the National Academy of Sciences 2021; 118(34): e2109229118.

14. Wu P, Liu F, Chang Z, et al. Assessing Asymptomatic, Presymptomatic, and Symptomatic Transmission Risk of Severe Acute Respiratory Syndrome Coronavirus 2. Clinical Infectious Diseases 2021; 73(6): e1314–e20.

15. Vaccine targets and forecasts. 2022. https://www.covid19data.com.au/vaccine-forecasts (accessed 9th February 2022).

16. Abdullah F, Myers J, Basu D, et al. Decreased severity of disease during the first global omicron variant covid-19 outbreak in a large hospital in tshwane, south africa. International Journal of Infectious Diseases 2022; 116: 38–42.

17. Maslo C, Friedland R, Toubkin M, Laubscher A, Akaloo T, Kama B. Characteristics and Outcomes of Hospitalized Patients in South Africa During the COVID-19 Omicron Wave Compared With Previous Waves. JAMA 2021.

18. Coronavirus (COVID-19) latest insights: Comparisons. 2021. https://www.ons.gov.uk/peoplepopulationandcommunity/healthandsocialcare/conditionsanddiseases/articles/coronaviruscovid19latestinsights/Overview (accessed 31st August 2021).

19. Coronavirus cases in Australia. 2022. https://covidlive.com.au (accessed 9th February 2022).

20. Miles J, Cramsie E. Queenslanders wanting COVID-19 test to ‘do the right thing’ or return to work left frustrated and confused. 11th Jan 2022 2022. https://www.abc.net.au/news/2022-01-06/qld-coronavirus-covid19-test-cant-get-one-now-what-need-work/100739784 (accessed 4th February 2022).

21. Covid border restrictions across Australia: where you can and can’t travel between states – and to New Zealand. September 3rd 2021 2021. https://www.theguardian.com/australia-news/2021/sep/03/covid-border-restrictions-australia-closures-bans-can-we-travel-victoria-vic-nsw-qld-queensland-wa-sa-act-nt-closures-where-you-can-not-go-fly-to-new-zealand-from-australia-nz (accessed February 28th 2022).

22. Australians to receive COVID-19 vaccine booster shot. Prime Minister of Australia; 2021.

23. Altarawneh H, Chemaitelly H, Tang P, et al. Protection afforded by prior infection against SARS-CoV-2 reinfection with the Omicron variant. medRxiv 2022: 2022.01.05.22268782.

24. Dayman I. South Australia opened its borders to COVID-19. So, what have we learned? 27th November 2021 2021. https://www.abc.net.au/news/2021-11-27/sa-borders-open-what-we-have-learnt-so-far/100655534 (accessed 1st March 2022).

25. Zillman S. What will life be like when Queensland reopens? How many COVID-19 cases will it see? What restrictions will be eased? 19th October 2021 2021. https://www.abc.net.au/news/2021-10-19/qld-coronavirus-covid-interstate-border-reopen-roadmap-christmas/100548346 (accessed 1st March 2022).

26. Brook B. Data shows Australians have reduced movement – but not to lockdown levels. Janurary 13th 2022 2022. https://www.news.com.au/technology/innovation/data-shows-australians-have-reduced-movement-but-not-to-lockdown-levels/news-story/9377d205e4ee778c901f301449bfd581 (accessed February 22nd 2022).

27. Estimates of the population for the UK, England and Wales, Scotland and Northern Ireland. 2021. https://www.ons.gov.uk/peoplepopulationandcommunity/populationandmigration/populationestimates/datasets/populationestimatesforukenglandandwalesscotlandandnorthernireland (accessed 31st August 2021).

28. Newcastle. 2013. https://quickstats.censusdata.abs.gov.au/census_services/getproduct/census/2011/quickstat/11103 (accessed 26th November 2019).

29. Lake Macquarie - East. 2013. https://quickstats.censusdata.abs.gov.au/census_services/getproduct/census/2011/quickstat/11101 (accessed 26th November 2019).

30. Litton E, Huckson S, Chavan S, et al. Increasing ICU capacity to accommodate higher demand during the COVID-19 pandemic. Med J Aust 2021; 215(11): 513–7.

31. Miles J, McKenna K, Hamilton-Smith L. Queensland health workers struggle with weight of ‘full-blown crisis’ of COVID-19’s Omicron surge. 13th January 2022 2022. https://www.abc.net.au/news/2022-01-13/qld-coronavirus-covid19-strain-on-health-sector-staff-omicron/100750292 (accessed 1st March 2022).

32. Coronavirus (COVID-19) Hospitalizations. 2022. https://ourworldindata.org/covid-hospitalizations (accessed February 22nd 2022).

33. Wen W, Chen C, Tang J, et al. Efficacy and safety of three new oral antiviral treatment (molnupiravir, fluvoxamine and Paxlovid) for COVID-19:a meta-analysis. Annals of Medicine 2022; 54(1): 516–23.

34. Takashita E, Kinoshita N, Yamayoshi S, et al. Efficacy of Antibodies and Antiviral Drugs against Covid-19 Omicron Variant. New England Journal of Medicine 2022.

35. COVID-19 vaccine surveillance report (week 7). In: Agency UHS, editor.; 2022. p. 57.

36. Sofonea MT, Roquebert B, Foulongne V, et al. From Delta to Omicron: analysing the SARS-CoV-2 epidemic in France using variant-specific screening tests (September 1 to December 18, 2021). medRxiv 2022: 2021.12.31.21268583.

37. Jalali N, Brustad HK, Frigessi A, et al. Increased household transmission and immune escape of the SARS-CoV-2 Omicron variant compared to the Delta variant: evidence from Norwegian contact tracing and vaccination data. medRxiv 2022: 2022.02.07.22270437.

38. Dunstan J. There’s no Victorian COVID-19 lockdown, but Melburnians are choosing to stay home anyway. 19th January 2022 2022. https://www.abc.net.au/news/2022-01-19/victoria-melbourne-covid-shadow-lockdown-cbd-mobility/100753496 (accessed 1st March 2022).

